# Incorporating clinical, genomic profiles and polygenic risk scores for the subtyping of depressive disorders

**DOI:** 10.1101/2023.03.01.23286610

**Authors:** Liangying Yin, Yuping Lin, Simon Sai-Yu Lui, Hon-Cheong So

## Abstract

Precise stratification of clinical patients into more homogeneous disease subgroups could address the heterogeneity of disease phenotypes and enhance our understanding on possible biological mechanisms and pathophysiology of more specified subtypes. This approach could promote individualized and effective prevention/intervention strategies. In the extant literature, subtyping of patients with depressive disorders (Dep) mainly utilized clinical features only. Genomics data could be useful subtyping features but advanced methods are needed for subtyping psychiatric entities such as depression. To solve this issue, we proposed a novel disease subtyping framework for complex diseases such as Dep. It combines brain structural features with genotype-predicted gene expression levels of relevant brain tissues as well as polygenic risk scores (PRS) of related disorders. It is able to classify patients into both clinically and biologically homogeneous subgroups, based on a multiview biclustering method. Moreover, causal inference was employed to identify causally relevant genes in different brain tissues to inform feature selection under the proposed framework. We verified the reliability of the subtyping model by internal and external validation. The calculated prediction strengths(PS) (average PS:0.896, min PS: 0.854) supported the robustness and generalizability of our proposed approach. External validation results demonstrated that our proposed approach could stratify Dep patients into subgroups with varied treatment responses and hospitalization risks. Besides, some subtype-defining genes in our study overlapped with several well-known susceptibility genes for depression and were involved in the pathophysiology for the disease. Encouragingly, many enriched drugs based on identified subtype-defining genes have been reported in previous studies to be effective in reducing depression-related symptoms.

## Introduction

The burden of depressive disorders (Dep) remains immense despite the extensive progress made by large-scale population studies, including genome-wide association studies and neuroimaging studies. According to the global burden of disease report, the depressive disorder ranked among the top 10 even 5 most disabling conditions across various age groups ^1^. Besides, the life expectancy for patients with depressive disorders is usually much shorter than the general population^1,2^. Precise classification of Dep patients diagnosed into more clinically and genetically homogeneous subgroups could facilitate our understanding on possible biological mechanisms and could address the issue of phenotype heterogeneity ^3,4^. Most importantly, it could shed light on the discovery of subtype-specific drug targets and promote individualized preventive and intervention strategies. In the extant literature, subtyping of patients with Dep mainly utilized clinical features only. The resultant Dep subtypes identified using clinical subtyping may not be efficient to have distinct and subgroup biological mechanisms. With the aid of genomic information, sophistic subtyping method can utilized complex data to stratify clinical patients into more biologically homogenous subgroups. Nevertheless, limited research have applied genomic data for stratification of Dep patients.

Over the past two decades, substantial efforts have been made to dissect the genetic architecture of depressive disorder (Dep) ^5,6^. Despite the substantial disorder-associated loci identified from GWAS, it remains very challenging to translate these findings into clinical usage, as many of the identified loci reside in non-coding regions, and only have minor effects on the studied disorder. Compared with SNP-based analysis, gene-based analysis is usually more biologically relevant and interpretable^3^. However, most gene-based studies mainly focused on univariate associations and did not qualify whether the identified trait-associated genes could actually contribute to the onset of disease, implicating possible spurious findings.

We proposed a novel framework for patient stratification by incorporating genotype-predicted gene expression profiles of relevant brain tissues with polygenic risk scores (PRS) and brain structural information (more specifically, volume of grey matter in different brain regions) by multi-view sparse bi-clustering analysis. PRS is a useful representation of the overall genetic predisposition of subjects to a particular disease ^7,8^. In this study, we proposed to use PRS of related diseases/traits as new features of our clustering analysis. Genes were selected based on a causal inference framework such that the most functionally relevant genes could be included for disease subtyping. Through imputing genetic variants to expression levels, the dimensionality will be significantly reduced. Since a causal inference method is employed to identify causally relevant genes for the studied disorder, the derived results will be more biologically relevant and easier to interpret. We hypothesized that the incorporation of brain structural and genetic information could help us identify more genetically coherent and clinically meaningful depression subtypes.

Our main contribution is presenting a novel framework for complex disease subtyping by leveraging genotype-predicted gene expression files of diverse relevant tissues, brain structural features as well as PRS of relevant disease. In this study, we incorporated brain structural features (the volume of grey matters in different brain regions) into our method. Compared with psychopathologies, neuroimaging characteristics are believed to be directly reflecting the results of biological mechanisms of the disease. Very few studies have attempted to utilize both brain structural features and genetic profiles for complex disease subtyping. Our work differs from existing studies by incorporating a causal algorithm of gene selection into a multi-view clustering framework for complex disease; we employ PC-simple algorithm ^9^ to identify causally relevant genes for our studies disease, then we apply a multi-view sparse clustering algorithm to stratify patients into different categories by incorporating brain structural features, PRS of relevant diseases and causally relevant genes in different tissues.*To our knowledge, we are the first to incorporate brain structural data, PRS of other diseases, as well as a causal algorithm of gene selection into a multi-view clustering framework for complex diseases*. Our method allows the genes to be selected based on a causal algorithm (i.e., PC-simple algorithm). Thus, the identified subtype-defining gene sets tend to be more functionally relevant to the underlying disease mechanisms.

## Method

### A novel stratification model

Our proposed framework could identify psychiatric disorders subtypes by incorporating genotype-predicted gene expression profiles of relevant brain tissues, variant-based PRSs of relevant disorders as well as brain structural information by a multi-view sparse biclustering method. PRS is a weighted sum of the risk allele count, with weights derived from log odds ratios or coefficients from regression. They are useful representations of the overall predisposition to a disease or trait. In this study, we incorporated PRS of relevant traits as new features of our clustering algorithm. More specifically, we employed “PRsice” ^12^ to calculate the variant based PRSs of relevant traits as new features for our clustering algorithm. By using a multi-view approach to clustering, it can uncover disease heterogeneity across different data views of patients (clinical and genetics).

In brief, our proposed framework comprised of 4 stages, i.e., data imputation, feature selection, disease subtyping and validation (as shown in Fig. 1). Next, we shall describe each step in greater detail below.

**Fig. 1.**
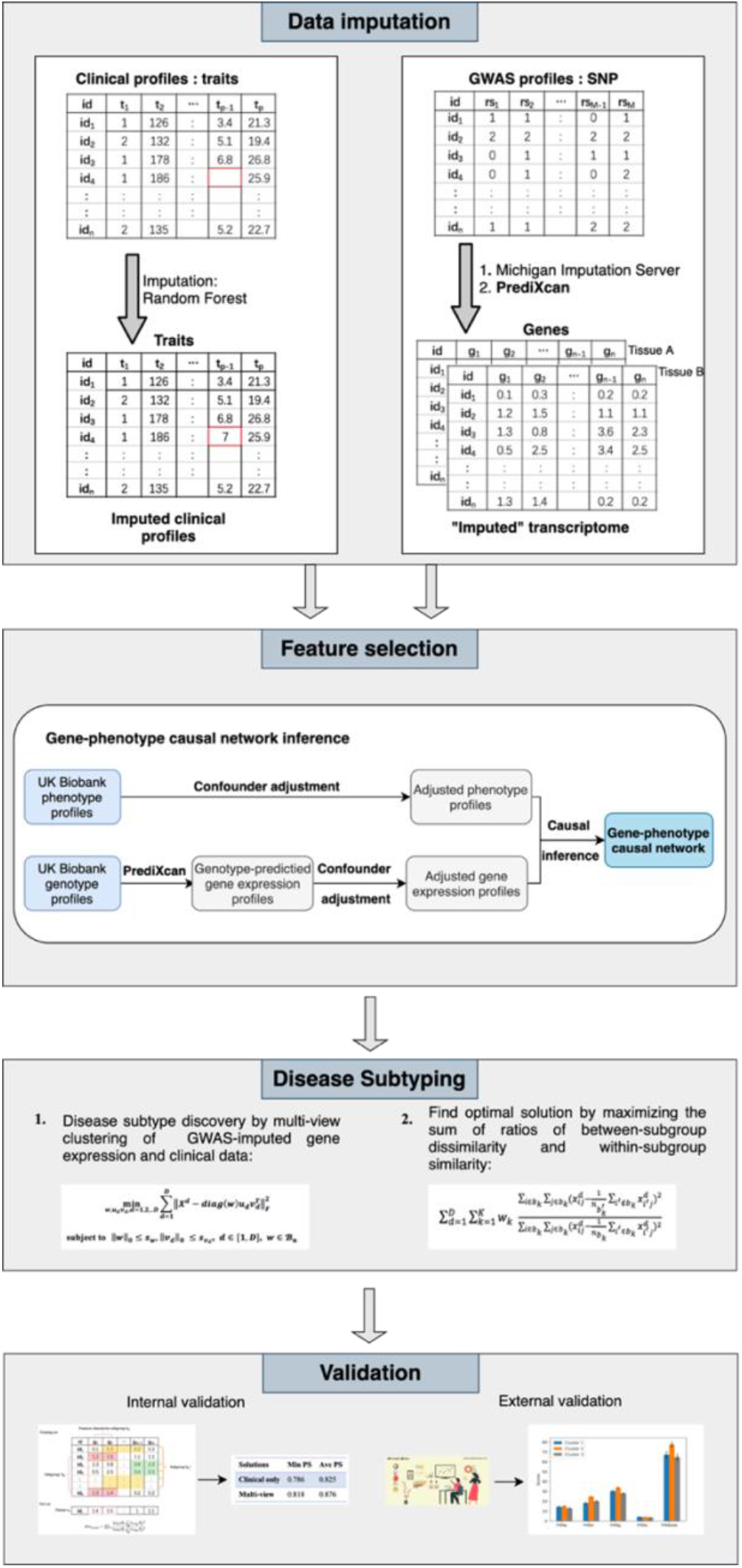
The workflow for the proposed invention in identifying disease subtypes

### Data imputation

Given that clustering analysis could not accommodate missing data, we employed imputation. Notably, different methods were used to impute missing clinical and genetic features. For clinical data, R package “missForest” ^13^ was employed to impute the missing data by a random forest algorithm. As for the estimation of transcriptomes from GWAS, we employed the “PrediXcan” developed by Gamazon et al. ^14^ to impute expression levels of relevant tissues form the genotype data. The algorithm first built elastic-net based prediction models with expression levels as the outcome from external reference dataset GTEx, which contained both genotype and expression data. Then, the developed prediction model was applied to new genotype data to “estimate” the expression levels of different tissues.

### Feature selection

In this study, we proposed to use a gene-phenotype causal network inference method to identify causally relevant genes for the disorder of interest. Fig. 1 shows the feature selection process. Confounder adjustments were separately performed for the disorder of interest and imputed expression profiles of the corresponding subjects. PC-simple algorithm ^9,15^ was employed to infer the causal relationships between genes and disorder, based on imputed expression profiles. In brief, PC-Simple can be regarded as a generalization of correlation screening that utilizes ordered independence screening algorithm to estimate the causal relationships between genes and studied phenotype. Let *X* = [*X*^1^,*X*^2^,…*X*^*p*^] be a *n* × *p* matrix of adjusted gene expression data for p genes, Y be a vector of the corresponding adjusted phenotype dataset for n subjects. Suppose Y is defined by a linear model of X, i.e.,:

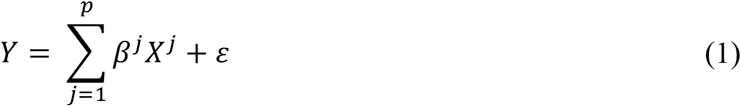

Where *ε* ∼*N*(0,∑) denotes the noise item, and it is independent of *X*^*j*^(*j* = 1,2,…*p*). For equation (1), we believe most or some of the coefficients *β*^*j*^ are zero, while the remaining are nonzero for the studied phenotype. Our goal was to uncover the active gene set *G* = {*j* = 1,2,…,*p*; *β*^*j*^ ≠ 0} with non-zero coefficients. Under the partial faithfulness assumption, we have:

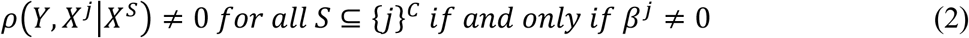

We could derive the active gene set through recursively performing partial correlation screening with increased order of conditional set based on (2) for all gene-phenotype pairs (*Y,X*^*j*^). Specifically, we first set the conditional set to null(*S* = ∅) and obtained the first candidate gene set with non-zero correlations with our studied phenotype. Then we sequentially increased the order of the conditional set to eliminate irrelevant genes until the candidate gene set did not vary anymore. The partial correlations for each gene-phenotype pair can be estimated as follows:

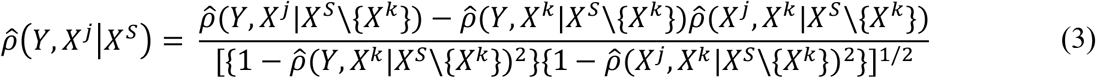

We tested whether calculated partial correlations by Fisher’s Z-transform, which can be expressed as follows:

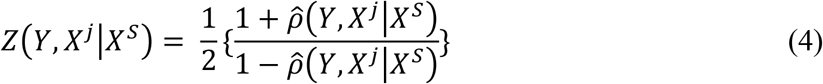

Buhlmann et al. ^9^ suggested to reject the null hypothesis 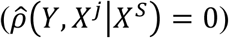 if the following inequation holds:

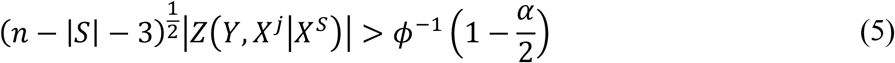

Where *ϕ* denotes standard normal cumulative distribution function, *α* denotes the significance level for the null hypothesis test. In this study, we set *α* = 0.05. To boost the computational efficiency, we set the maximum order for partial correlation screening to 3. All genes survived the 3-order partial correlation screening were regarded as directly causal genes for the studied phenotype. After deriving the tissue-specific gene-phenotype causal graph for the studied phenotype, we could distinguish the directly causal genes from other ones. The identified gene-phenotype causal network could be utilized to inform the feature selection for the subsequent disease subtyping process.

### Disease subtyping

For disease subtype discovery, we employed an extension of the biclustering algorithm in ref ^16^. In brief, we performed biclustering by matrix decomposition. Suppose *X*^*d*^ is a *n* × *m*_*d*_ data matrix from the clinical or genetic view of patients, where n is the sample size, *d* denotes the index of ‘view’ to be modeled and *m*_*d*_ is the number of features in the dth view. For example, if one models clinical and genotype-predicted expressions in one tissue, there will be two views. It is possible to extend the approach to more than 2 views, for example based on expression in different tissues or using other (preferably gene-based) ‘omics’ profiles. It is worth emphasizing that due to heterogeneity of patients, the pathophysiology (e.g. genetic pathways) underlying the disease may be different for different subgroups of patients. Using a biclustering algorithm, each bicluster can be characterized by different sets of gene features; in other words, we allow different genes to be involved in the disease for different subgroups of patients. This adds flexibility to our model and is an important advantage compared to ordinary cluster approaches. Subgroups of patients can be simultaneously derived by performing a sparse rank one approximation on the original matrices *X*^*d*^ (*d* = 1,2,.. *D*, indicating data matrices from different views that characterize the same set of patients), i.e.,

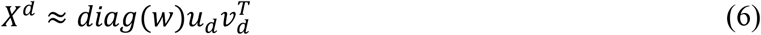

where *w* is a binary vector of size *n*, serving as a common factor that forces different views of data to agree on the same grouping of patients. *diag*(*w*) is a diagonal matrix of size *n* × *n* with diagonal entries equal to *w. u*_*d*_ of size *n* and *v*_*d*_ of size *m*_*d*_ are the rank-one approximations of *X*^*d*^ respectively. Rows in *X*^*d*^ corresponding to the non-zero entries of *diag*(*w*) form the row subgroups, and columns in *v*_*d*_ form the column subgroups (a.k.a., sub-feature groups) in different views. Subgroups of patients based on different views of data can be derived by solving the following optimization problem:

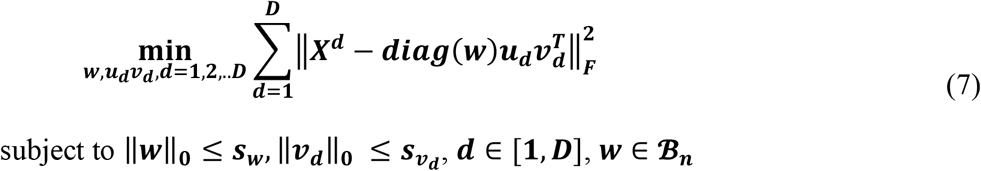

where *s*_*w*_ and 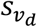’s are hyper-parameters that need to be predetermined to enforce sparsity of *w* and *v*_*d*_ ‘s, i.e., the number of patients 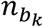 and number of selected features 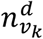 in each subgroup of the corresponding data view. *D* is the number of data views incorporated for clustering and *B*_*n*_ is the set that contains all possible binary vectors of length *n*. To obtain subsequent subgroups, we need to first update the data matrices by excluding previously identified patients, then solve Eq. (7). Our proposed approach is capable of selecting features during the clustering process, however, we need to predetermine the number of selected features in each data view. In this invention, we follow the suggestions given by the original authors. More specifically, the number of selected features 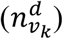 in each data view will be set to the number where the accumulated variance in PCA of *X*^*d*^ was over 90%. The algorithm requires the number and size of subgroups to be specified beforehand. We consider a value range of 2 to 6 for the number of subgroups and the minimum number of subjects in each subgroup (min 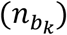) was set to 20. Suppose the number of subgroups is k, the size of each subgroup will be firstly set to a value roughly equals to *n*/*k*. Then, all combinations by adding or subtracting min 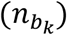 in each subgroup will be tried. A grid search approach will be employed to determine the optimal solution. An evaluation metric is required to find the optimal solution. One of the most used metrics is mean squared residue (MSR). However, it only assesses the homogeneity within each subgroup and does not consider the heterogeneity between different subgroups. For well-separated subgroups, patients within the same subgroups should be highly homogenous while patients belonging to different subgroups should be highly heterogeneous. In view of this, we employed the sum of ratios of between biclusters distance and within biclusters distance (BBD/WBD) proposed by Yin et al. ^3^ as the evaluation metric to identify the optimal solution.

### Validation

We employed external and internal validations (as shown in Fig. 1) to our resultant subtypes. Regarding external validation, this method was used when external data of the disease outcomes (as validating rather than clustering variables) was available. Regarding internal validation, this method was used only when external validation was not feasible. Specifically, internal validation utilized the extended “prediction strength” (PS) method developed by Yin et al. ^3^, which would be applicable for multi-view clustering analysis as validation of identified subgroups. First, we split the sample into a “training set” and another “testing set”, and then evaluated whether the disease subtyping model derived from the training set could “predicts” the actual disease subgroups derived from the training set alone. In essence, it measured how well the “predicted” co-memberships (based on the model derived from training set) in the testing set could matche with actually performing cluster analysis on it. In this study, we calculated both the “min PS” and “ave PS” to evaluate the performance of our proposed method. The “min PS” and “ave PS” respectively measured the lowest and average proportion of co-memberships among all identified subtypes, as follow:

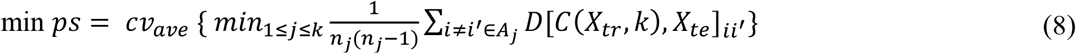

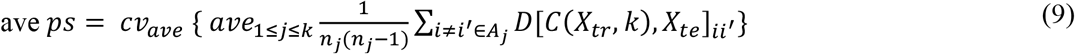

Here *C*(*X*_*tr*_,*k*) indicates the clustering operation on the training set with *k* subgroups. *D*[*C*(*X*_*tr*_,*k*),*X*_*te*_]_*ii*_*′* denotes the co-membership for subjects *i* and *i*^*′*^ in subgroup *A*_*j*_. *n*_*j*_ is the number of subjects in subgroup *A*_*j*_. *cv*_*ave*_ refers to the average of all cross validation folds. Tibshirani et al. ^17^ suggested that a PS of 0.8 or above indicates reasonably good predictions strength.

### Further analyses

To further validate the identified subgroups, we examined whether genes selected by our proposed approach were enriched for GWAS hit of depression. Specifically, the GWAS summary statistics for depression was first converted to gene-based statistics by FASTBAT ^18^, then we tested whether genes selected by our framework had lower p-value than non-selected one.

In addition, we extracted genes selected by our method to figure out the genetic underpinning of each identified depression subtype. Pathway analyses were also conducted on “ConsensumPathDB” ^19,20^ to further exploring the pathophysiology and biological mechanisms underlying each subtype. In other words, we performed over-representation analyses on the subtype-defining gene sets identified in each genetic view by the hypergeometric test. Furthermore, we conducted drug enrichment analyses on ‘Enrichr’ ^21^ to identify potentially conducive drugs for each subtype.

### Application to depression patients

We applied our disease subtyping model to depression-affected subjects collected in the UK biobank (UKBB). Here, depression is defined by a combination of ICD-10 coded and self-reported disease. Since high missing rate may affect the imputation accuracy of the applied imputation method, we only keep patients with a comparably lower clinical missing rate. Some researchers believed that 10% is a reasonable missingness cutoff with satisfactory imputation accuracy 13,14. Following this, we only keep patients with a clinical missing rate less than 10%. More specifically, all depression patients with available brain structural features whose missing rates were less than 10% were reserved for further analysis. We imputed the clinical data by R package “missForest” ^13^ with default setting. Besides, we estimated the expression levels for cortex, frontal cortex, nucleus accumbens basal ganglia and putamen basal ganglia by “PrediXcan” ^14^ for all subjects with available genotypes. Evidence suggests the intensive involvement of these brain tissues in the pathophysiology of depression ^22-27^. Thus, we incorporated these 4 tissues as the genetic views of our clustering algorithm. Apart from this, we also calculate the PRSs of related neuropsychiatric traits. The traits for constructing PRS included autism spectrum disorders (ASD; N=46,350) ^28^, attention deficit hyperactivity disorder (ADHD; N=53,293) ^29^, schizophrenia (SCZ; N=105,318) ^30^, bipolar disorder (BP; N=41,653) ^31^, major depressive disorder (MDD; N=500,199) ^32^ and post-traumatic stress disorder (PTSD; N=200,000) ^33^. GWAS summary statistics were downloaded from Psychiatric Genomics Consortium (PGC) (https://www.med.unc.edu/pgc) and The Integrative Psychiatric Research project (iPSYCH). Before the standard PRS analysis, LD-clumping was required. In this application, we performed LD-clumping at *r*^2^ = 0.1 within a distance of 1000 Kb ^34^. PRS was generated by PRsice with a P-value threshold of 0.1. We incorporated these 6 PRSs as clinical features. In total, we included 139 brain structural features (volume of grey matter in different brain regions) and 6 PRSs of related neuropsychiatric disorders as input features in the clinical view.

## Results

### Subtyping results

Based on the definition of depression, we extracted 28,335 depression-affected subjects as cases and 28,5921 as controls from UKBB as the input for the causal inference-based feature selection. To avoid possible biases introduced by population structure, we adjusted the predicted tissue-specific expression profiles and phenotype data by the top 10 principal components (PCs) of the corresponding genotype dataset first. Then we used the corrected input data to identify causally relevant genes for depression in each tissue. We respectively identified 108, 101, 94 and 76 genes for cortex, frontal cortex, nucleus accumbens basal ganglia and putamen basal ganglia. These genes were persevered as input in the corresponding genetic views for the subtyping of 352 depression-affected patients.

In this study, we incorporated 5 different views for the subtyping of depression-affected patients, one clinical view with 139 brain structural features and PRS of 6 related disorders, and 4 genetic views with predicted gene expression profiles for causally relevant genes. As mentioned earlier, PCA was employed to determine the number of selected features in each data view. Table 1 lists the number of selected features in the corresponding data view. The best performance was achieved when the depression-affected subjects were stratified into 2 different subgroups with 20 and 332 subjects in each subgroup. For clinical features, 63 out of 145 features were selected as subtype-defining features (see Table 1), and all of them were brain structural features (Table 2). From the perspective of algorithm, features selected by the algorithm usually are more relevant and informative than the non-selected ones. Table 2 summarizes the comparison results of subtype-defining clinical features. We observed significant differences on selected features between two identified subgroups (Fig. 2). Notably, among the selected subtype-defining brain structural features, most were shared in two subtypes. To be more specific, 51 out of 63 clinical features were overlapped between to identified subtypes, while the remaining 12 were subtype specific.

**Table 1.** The number of selected features in each data view

| Data view | No. of selected features |
| --- | --- |
| Clinical | 63 |
| Cortex | 80 |
| Frontal cortex | 76 |
| Nucleus accumbens basal ganglia | 71 |
| Putamen basal ganglia | 59 |

**Table 2.** Comparison results for subtype-defining clinical features

| Subtype 1 |  | Subtype 2 |  |
| --- | --- | --- | --- |
| Features | P-value | Features | P-value |
| Volume of grey matter in Frontal Pole (left) | 5.89E-08 | Volume of grey matter in Frontal Pole (left) | 5.89E-08 |
| Volume of grey matter in Frontal Pole (right) | 1.30E-05 | Volume of grey matter in Frontal Pole (right) | 1.30E-05 |
| Volume of grey matter in Insular Cortex (left) | 1.11E-04 | Volume of grey matter in Insular Cortex (left) | 1.11E-04 |
| Volume of grey matter in Insular Cortex (right) | 5.02E-04 | Volume of grey matter in Insular Cortex (right) | 5.02E-04 |
| Volume of grey matter in Superior Frontal Gyrus (left) | 3.11E-04 | Volume of grey matter in Superior Frontal Gyrus (left) | 3.11E-04 |
| Volume of grey matter in Superior Frontal Gyrus (right) | 4.66E-03 | Volume of grey matter in Superior Frontal Gyrus (right) | 4.66E-03 |
| Volume of grey matter in Middle Frontal Gyrus (left) | 2.35E-04 | Volume of grey matter in Middle Frontal Gyrus (left) | 2.35E-04 |
| Volume of grey matter in Middle Frontal Gyrus (right) | 3.56E-08 | Volume of grey matter in Middle Frontal Gyrus (right) | 3.56E-08 |
| Volume of grey matter in Precentral Gyrus (left) | 2.85E-03 | Volume of grey matter in Inferior Frontal Gyrus, pars opercularis (right) | 1.25E-04 |
| Volume of grey matter in Precentral Gyrus (right) | 1.40E-04 | Volume of grey matter in Precentral Gyrus (left) | 2.85E-03 |
| Volume of grey matter in Temporal Pole (left) | 2.39E-04 | Volume of grey matter in Precentral Gyrus (right) | 1.40E-04 |
| Volume of grey matter in Temporal Pole (right) | 1.61E-04 | Volume of grey matter in Temporal Pole (left) | 2.39E-04 |
| Volume of grey matter in Superior Temporal Gyrus, anterior division (right) | 2.77E-03 | Volume of grey matter in Temporal Pole (right) | 1.61E-04 |
| Volume of grey matter in Superior Temporal Gyrus, posterior division (right) | 2.26E-05 | Volume of grey matter in Superior Temporal Gyrus, posterior division (right) | 2.26E-05 |
| Volume of grey matter in Middle Temporal Gyrus, anterior division (left) | 2.62E-04 | Volume of grey matter in Middle Temporal Gyrus, anterior division (left) | 2.62E-04 |
| Volume of grey matter in Middle Temporal Gyrus, anterior division (right) | 7.79E-03 | Volume of grey matter in Middle Temporal Gyrus, posterior division (left) | 1.24E-04 |
| Volume of grey matter in Middle Temporal Gyrus, posterior division (left) | 1.24E-04 | Volume of grey matter in Middle Temporal Gyrus, posterior division (right) | 2.27E-04 |
| Volume of grey matter in Middle Temporal Gyrus, posterior division (right) | 2.27E-04 | Volume of grey matter in Postcentral Gyrus (left) | 2.91E-03 |
| Volume of grey matter in Postcentral Gyrus (left) | 2.91E-03 | Volume of grey matter in Postcentral Gyrus (right) | 4.03E-05 |
| Volume of grey matter in Postcentral Gyrus (right) | 4.03E-05 | Volume of grey matter in Lateral Occipital Cortex, superior division (left) | 5.54E-05 |
| Volume of grey matter in Supramarginal Gyrus, posterior division (right) | 1.37E-04 | Volume of grey matter in Lateral Occipital Cortex, superior division (right) | 1.23E-03 |
| Volume of grey matter in Lateral Occipital Cortex, superior division (left) | 5.54E-05 | Volume of grey matter in Lateral Occipital Cortex, inferior division (left) | 2.95E-06 |
| Volume of grey matter in Lateral Occipital Cortex, inferior division (right) | 2.70E-05 | Volume of grey matter in Lateral Occipital Cortex, inferior division (right) | 2.70E-05 |
| Volume of grey matter in Frontal Medial Cortex (left) | 9.62E-04 | Volume of grey matter in Subcallosal Cortex (left) | 7.64E-04 |
| Volume of grey matter in Frontal Medial Cortex (right) | 2.61E-03 | Volume of grey matter in Subcallosal Cortex (right) | 1.63E-03 |
| Volume of grey matter in Subcallosal Cortex (left) | 7.64E-04 | Volume of grey matter in Paracingulate Gyrus (left) | 2.68E-05 |
| Volume of grey matter in Subcallosal Cortex (right) | 1.63E-03 | Volume of grey matter in Paracingulate Gyrus (right) | 3.51E-04 |
| Volume of grey matter in Paracingulate Gyrus (left) | 2.68E-05 | Volume of grey matter in Cingulate Gyrus, posterior division (left) | 3.41E-05 |
| Volume of grey matter in Paracingulate Gyrus (right) | 3.51E-04 | Volume of grey matter in Cingulate Gyrus, posterior division (right) | 6.63E-05 |
| Volume of grey matter in Cingulate Gyrus, anterior division (right) | 1.98E-03 | Volume of grey matter in Precuneous Cortex (left) | 2.29E-05 |
| Volume of grey matter in Cingulate Gyrus, posterior division (left) | 3.41E-05 | Volume of grey matter in Precuneous Cortex (right) | 3.65E-05 |
| Volume of grey matter in Cingulate Gyrus, posterior division (right) | 6.63E-05 | Volume of grey matter in Cuneal Cortex (right) | 7.68E-04 |
| Volume of grey matter in Precuneous Cortex (left) | 2.29E-05 | Volume of grey matter in Frontal Orbital Cortex (left) | 1.81E-07 |
| Volume of grey matter in Precuneous Cortex (right) | 3.65E-05 | Volume of grey matter in Frontal Orbital Cortex (right) | 1.30E-05 |
| Volume of grey matter in Cuneal Cortex (left) | 8.96E-03 | Volume of grey matter in Parahippocampal Gyrus, anterior division (left) | 1.94E-03 |
| Volume of grey matter in Cuneal Cortex (right) | 7.68E-04 | Volume of grey matter in Parahippocampal Gyrus, anterior division (right) | 9.30E-03 |
| Volume of grey matter in Frontal Orbital Cortex (left) | 1.81E-07 | Volume of grey matter in Lingual Gyrus (left) | 3.75E-04 |
| Volume of grey matter in Frontal Orbital Cortex (right) | 1.30E-05 | Volume of grey matter in Lingual Gyrus (right) | 1.98E-04 |
| Volume of grey matter in Lingual Gyrus (left) | 3.75E-04 | Volume of grey matter in Temporal Fusiform Cortex, anterior division (left) | 1.78E-03 |
| Volume of grey matter in Lingual Gyrus (right) | 1.98E-04 | Volume of grey matter in Temporal Fusiform Cortex, anterior division (right) | 7.86E-04 |
| Volume of grey matter in Temporal Fusiform Cortex, posterior division (left) | 6.63E-03 | Volume of grey matter in Temporal Fusiform Cortex, posterior division (left) | 6.63E-03 |
| Volume of grey matter in Temporal Fusiform Cortex, posterior division (right) | 1.29E-02 | Volume of grey matter in Temporal Fusiform Cortex, posterior division (right) | 1.29E-02 |
| Volume of grey matter in Occipital Fusiform Gyrus (right) | 3.42E-03 | Volume of grey matter in Occipital Fusiform Gyrus (right) | 3.42E-03 |
| Volume of grey matter in Central Opercular Cortex (left) | 1.14E-05 | Volume of grey matter in Frontal Operculum Cortex (left) | 1.17E-03 |
| Volume of grey matter in Central Opercular Cortex (right) | 4.64E-05 | Volume of grey matter in Frontal Operculum Cortex (right) | 7.39E-04 |
| Volume of grey matter in Parietal Operculum Cortex (right) | 5.08E-05 | Volume of grey matter in Central Opercular Cortex (left) | 1.14E-05 |
| Volume of grey matter in Planum Polare (right) | 1.05E-04 | Volume of grey matter in Central Opercular Cortex (right) | 4.64E-05 |
| Volume of grey matter in Heschl's Gyrus (includes H1 and H2) (left) | 1.63E-03 | Volume of grey matter in Parietal Operculum Cortex (left) | 6.00E-04 |
| Volume of grey matter in Heschl's Gyrus (includes H1 and H2) (right) | 6.25E-04 | Volume of grey matter in Parietal Operculum Cortex (right) | 5.08E-05 |
| Volume of grey matter in Planum Temporale (left) | 1.29E-02 | Volume of grey matter in Planum Polare (left) | 5.24E-05 |
| Volume of grey matter in Planum Temporale (right) | 5.08E-05 | Volume of grey matter in Planum Polare (right) | 1.05E-04 |
| Volume of grey matter in Supracalcarine Cortex (left) | 1.54E-03 | Volume of grey matter in Heschl's Gyrus (includes H1 and H2) (left) | 1.63E-03 |
| Volume of grey matter in Supracalcarine Cortex (right) | 5.60E-06 | Volume of grey matter in Heschl's Gyrus (includes H1 and H2) (right) | 6.25E-04 |
| Volume of grey matter in Occipital Pole (left) | 3.96E-05 | Volume of grey matter in Planum Temporale (left) | 1.29E-02 |
| Volume of grey matter in Occipital Pole (right) | 3.53E-04 | Volume of grey matter in Planum Temporale (right) | 5.08E-05 |
| Volume of grey matter in Thalamus (left) | 5.66E-02 | Volume of grey matter in Supracalcarine Cortex (left) | 1.54E-03 |
| Volume of grey matter in Thalamus (right) | 3.27E-02 | Volume of grey matter in Supracalcarine Cortex (right) | 5.60E-06 |
| Volume of grey matter in Hippocampus (left) | 5.17E-04 | Volume of grey matter in Occipital Pole (left) | 3.96E-05 |
| Volume of grey matter in Hippocampus (right) | 5.08E-03 | Volume of grey matter in Occipital Pole (right) | 3.53E-04 |
| Volume of grey matter in Amygdala (right) | 6.36E-06 | Volume of grey matter in Hippocampus (left) | 5.17E-04 |
| Volume of grey matter in V Cerebellum (left) | 3.67E-02 | Volume of grey matter in Hippocampus (right) | 5.08E-03 |
| Volume of grey matter in V Cerebellum (right) | 3.03E-02 | Volume of grey matter in Amygdala (left) | 1.79E-04 |
| Volume of grey matter in VI Cerebellum (right) | 3.19E-02 | Volume of grey matter in Amygdala (right) | 6.36E-06 |

**Fig. 2.** Comparison subtype-defining clinical features for depression-affected patients

Using the UKBB, we gathered prognoses-related variables for these depression-affected subjects. More specifically, we extracted the treatment resistance status and admission frequency of corresponding patients from the general practitioner (GP) records to evaluate our identified depression subtypes. In this study, treatment resistance depression (TRD) was defined as depression-affected patients who tried at least two different antidepressant drugs. Fabbrri et al. ^35^ suggested that the time interval between two drugs should be no longer than 14 weeks and each drug should be prescribed for at least 6 weeks.

Since the medication records are not available for all subjects in the UKBB, we only gathered the treatment resistance status for 292 patients. When comparing the differences in TRD between the two derived subgroups, patients with missing values were excluded. Fisher exact test was performed to examine whether there exist significant differences between the two derived subgroups regarding the missing rate. Notably, the results did not find any significant differences between the two derived subgroups in accessibility of patient records for the TRD status(p-value=0.356). We compared the differences by a regression model and significant differences in TRD were observed between the two derived subgroups with a p-value of 0.049 (Fig. 3).

**Fig. 3.**
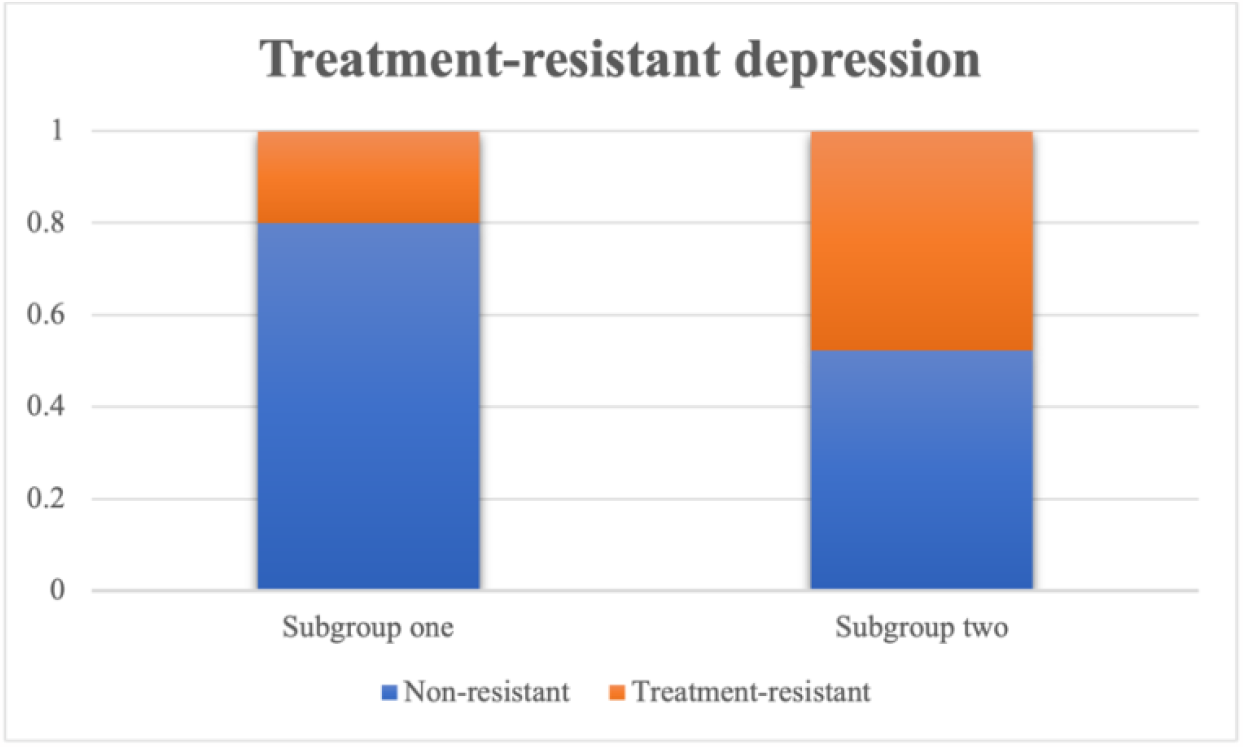
Comparison of TRD status by subgroups for depression-affected patients

Moreover, we compared the differences in depression-caused admission frequencies across the two derived subgroups by the Wilcoxon signed rank test. In line with TRD, subjects with missing values were excluded. The admission frequency records for 307 patients were available for further analysis. Fisher exact test did not find any no significant difference was observed in the availability of patients’ records for depression-related admission frequencies(p-value=0.730). Again, a significant difference between the two subgroups was observed with a p-value of 0.033.

Furthermore, we computed the extended prediction strength (PS) of our identified solution and obtained a minimum PS of 0.854 and an average PS of 0.896 (Table 3). A PS of >=0.8 suggested the stability of the tested model and generalizability to new datasets. Therefore, we could conclude that our proposed method is reliable and stable in revealing depression subtypes. Moreover, we compared our method with the conventional only clinical variable-based disease subtyping (Table). Table 3 shows that our proposed method could achieve higher “min PS” and “ave PS” than the clinical variable-only subtyping method despite its high complexity.

**Table 3.** Comparison of extended PS derived from different solutions

| Solutions | Min PS | Ave PS |
| --- | --- | --- |
| **Clinical only** | 0.786 | 0.825 |
| **Multi-view** | 0.854 | 0.896 |

### Results of further analyses

We downloaded the GWAS summary statistic for depression from PGC to examine whether our selected genes were enriched for the corresponding GWAS hit. Table 4 shows the enrichment analysis results. As expected, the genes picked up by our proposed framework were indeed enriched for known genes for depression. More specifically, subtype-defining genes for nucleus accumbens basal ganglia and putamen basal ganglia were significantly enriched for GWAS hit of depression.

**Table 4.** Enrichment analysis results for GWAS hits of depression

| Subgroups | P(Cortex) | P(Frontal cortex) | P(Nucleus accumbens basal ganglia) | P(Putamen basal ganglia) |
| --- | --- | --- | --- | --- |
| **One** | 0.3726 | 0.5027 | **0.0343** | **0.0547** |
| **Two** | 0.1436 | 0.5644 | **0.038** | **0.0112** |

Table S1 demonstrates the subtype-defining gene sets identified by our proposed framework. While many subtype-defining genes appeared in both depression subtypes, others were subtype-specific. Most importantly, many were well-known susceptibility genes for depression or involved in the related pathophysiological process. For example, ROGDI, NALCN, ITIH4 were selected by our algorithm as subtype-defining genes in both subtypes. A recent study by Gonda et al.^36^ suggested the influence of ROGD1 on anxious temperaments through its regulation of leucine zipper protein encoding in the brain. Studies demonstrate the connections between NALCN and various psychiatric disorders, including depression, bipolar, and so on. Cochet-Bissuel et al. ^37^ revealed its critical role in regulating neuronal excitability, and the alternation of NACLN channelosome may lead to the onset of depression-related symptoms. SNX19 was identified as a subtype-defining gene for subtype 2 only. According to Ma et al. ^38^, it is intimately involved in the initiation of the molecular mechanism of risk for schizophrenia, a highly comorbid disorder for depression. For more details, please refer to Table S1.

Table S2 lists the top enriched pathways that characterize each disease subtype. Numerous enriched pathways were involved in depression or related pathophysiology. Again, some enriched pathways were shared among two identified subtypes while others were subtype-specific. Here we highlighted a few pathways that may be of interest. Regulation of PTEN stability and activity, tyrosine metabolism were found to be significantly enriched for depression patients belonging to subtype 1. Wang et al. conducted an experiment on mice and found that regulation of PTEN stability and activity could lead to an increase in depression-related behaviors ^39^. Tyrosine metabolism was confirmed to be intimately related to anhedonia, a core symptom of depression, by Bekhbat et al ^40^. They also suggested that this pathway defined a subtype of depression. As for subtype 2, some of the significantly enriched pathways included PIP3 activates AKT signaling, prostaglandin (PG) synthesis and regulation and Vitamin D Receptor Pathway. A previous study by Matsuda et al. ^41^ suggested that PIP3 activates AKT signaling pathway plays a critical role in the survival of various neuron cells, which may evoke depression-related behaviors. Chu et al. ^42^ reported that PG synthesis and regulation could lead to the onset of depression through downregulating PG D2 levels in the plasma. For more details about other enriched pathways, please refer to Table S2.

Table S3 summarizes the enriched drugs for each identified depression subtype. Many were proven to be effective in reversing depression-related symptoms or behaviors. For example, apigenin, kaempferol and ouabain etc. A previous study by Weng at al. ^43^ found that apigenin could reverse depression-like behaviors in mice. Silva dos Santos et al. ^44^ implicated that kaempferol has a multipotential neuroprotective effect on depression. For more details on the corresponding enriched drugs, please refer to Table S3.

## Discussion

In this proof-of-concept study, we proposed a novel framework to identify depression subtypes by incorporating both clinical and genetic information using a multi-view biclustering method. We demonstrated the validity of our proposed framework by applying it to depression-affected subjects extracted from the UKBB. Two different depression subtypes with significant differences in the treatment resistance status and hospitalization frequencies could be identified. Internal validation result (PS) also revealed the stability of our proposed method in revealing depression subtypes and the generalizability to new datasets. Furthermore, the subtype-defining genes were significantly enriched for GWAS hits for depression. More encouragingly, many enriched drugs based on identified subtype-defining genes were proven to be effective in reversing depression-related symptoms or behaviors. We believe this proof-of-concept study shall be generalizable to a larger sample size.

This study has several advantages. A key advantage is that it’s the first study attempting to classify depression patients into homogeneous subgroups by incorporating both clinical and genetic information using a multi-view biclustering method. It combined brain structural features with PRSs of related disorders as well as genotype-imputed expression profiles of related brain tissues for the subtyping of depression. We note that some previous studies used PRS for the prediction of psychiatric disorder risk. However, none of them combined this with other clinical and genetic information for the subtyping of psychiatric disorders, especially depression. Secondly, causal inference was employed to identify causally relevant genes in different brain tissues to inform the feature selection process. Thus, subgroup-selected genes from this study are more likely to be clinically relevant and interpretable for the involved disease mechanisms for depression patients. To our knowledge, no previous works have employed a causal inference approach to select genes for clustering a complex disease/trait. Thirdly, our proposed framework allowed transcriptomes from different tissues to be modelled, which dramatically improved the flexibility of our method. Besides, it’s less invasive and more cost-effective than access to raw gene expression data. Since the gene expression levels were predicted from the genotypes, they were unlikely to be confounded by other factors, such as medication usage. Even though the proposed approach is applied to stratify depression patients into different subtypes, it is generalizable and could be used to subtype other psychiatric and neurological disorders.

Here we highlight a few related research. One related approach is to construct the polygenic risk score (PRS) for each subject first, then use the PRS to stratify subjects. There is a fundamental difference between our proposed method and the PRS. PRS aims to estimate the overall genetic liability to a particular disease^10^. However, it is not designed to uncover subtype-defining gene set. It’s more efficient in distinguishing cases from controls instead of stratifying confirmed cased into different subgroups. It’s very challenging to stratify patients solely by PRS as depression is a highly heterogeneous disorder. Patients with similar overall genetic liability may have distinct genetic basis^11^.

Previously, Yin et al.^3,4^ proposed to incorporate clinical and genetic data to subtype schizophrenia patients by a multi-view biclustering method. Our method is different from this work in several ways. First, *brain structural features* were employed as part of the features for the subtyping of depression patients while the previous work used well-defined clinical features for disease subtyping. The brain structural features used here carry useful information related to the underlying disease mechanisms that probably not reflected in well-defined clinical features. Compared with clinical symptoms, brain structural features can be more objectively assessed and are more directly linked to the underlying disease biology. In addition, because these structural features can be directly extracted from magnetic resonance imaging (MRI), and clinicians are not necessarily involved in this feature extraction process, which can save the cost/time of assessment by psychiatrists. Different from the previous work, the selection of subgroup-relevant clinical features was totally data-driven in this study covering hundreds of features instead of restricting to a small set of predefined clinical features (around 10) in the prior papers. This may lead to the discovery of new disease biomarkers. Besides, *PRS, which reflects the overall genetic predisposition of patients to related disorders, were also incorporated for the subtyping of depression patients in this invention*. To our knowledge, this is the first study that combine PRSs of related disorders and brain structural features as well as genotype-predicted expression profiles for the subtyping of depression patients, and for any complex diseases. In the above mentioned two publications, PRSs of related disorders were <u>not</u> included for the subtyping of patients. Moreover, *causal inference was employed to identify causally-relevant genes* to inform gene selection under the proposed framework. In the prior work, selection of genes was **not** based on any causal discovery methods. Even though genes selected from their frameworks may be associated with the identified subgroups, they may be **confounded** by other genes (i.e. the association may be spurious). Our proposed approach could differentiate causal genes from non-causal ones. Thus, subgroup-specific genes selected from our invention are more likely to be clinically relevant and interpretable to explain the contributing disease mechanisms of the studied disorder.

Several limitations of this study should be borne in mind. First, the sample size of the depression-affected patients dataset was modest. Even though it’s hard to collect data with both available brain structural features and genotypes, larger sample size will add diversity and be better in demonstrating whether our proposed method adapts well to a highly variable dataset. We believe this proof-of-concept study shall be generalizable to a larger sample size. Second, we failed to replicate our proposed clustering model in the independent datasets, because such additional datasets with brain structural features and genotype information were not available. Instead, we demonstrated the validity and feasibility of our proposed approach by extended prediction strength and external validation. Third, we had a very limited access to different outcome-related variables for validating our subgroups. Future research may have better access to outcome-related variables, because more patient-related data would be released in the UKBB. To conclude, we have proposed a novel disease subtyping model, which was capable of identifying depression subtypes by utilizing genotype-predicted expression levels of relevant brain tissues and brain structural information (more specifically, the volume of grey matter in different brain regions) as well as PRS of other diseases. Genes were selected based on a causal inference framework such that the most functionally relevant genes could be included for disease subtyping. Our proposed approach has opened a new avenue for exploring GWAS, brain structural features and PRS for the subtyping of psychiatric/neurological disorders. We believe this is a valuable endeavor to exploit the usage of causal inference for translational medicine.

## Supporting information

https://docs.google.com/spreadsheets/d/13okf_gSIFYt2r_qCIoFOMU9S6VdPuLjA/edit?usp=share_link&ouid=108055343734926668816&rtpof=true&sd=true

https://docs.google.com/spreadsheets/d/12bwz3-Wquk0FM-ZaaHvnqgvHada0GkLW/edit?usp=share_link&ouid=108055343734926668816&rtpof=true&sd=true

https://docs.google.com/spreadsheets/d/1lGvhsV__h34hKPwxsh3a_zkg404qfvhL/edit?usp=share_link&ouid=108055343734926668816&rtpof=true&sd=true

## Data Availability

All data produced in the present study are available upon reasonable request to the authors.

## Acknowledgements

This work was supported partially by an Innovation and Technology Fund (ITS/113/19), a National Natural Science Foundation China grant (81971706), the Lo Kwee Seong Biomedical Research Fund from The Chinese University of Hong Kong and the KIZ-CUHK Joint Laboratory of Bioresources and Molecular Research of Common Diseases, Kunming Institute of Zoology and The Chinese University of Hong Kong, China.

